# Control with uncertain data of socially structured compartmental epidemic models

**DOI:** 10.1101/2020.04.27.20081885

**Authors:** Giacomo Albi, Lorenzo Pareschi, Mattia Zanella

## Abstract

The adoption of containment measures to reduce the amplitude of the epidemic peak is a key aspect in tackling the rapid spread of an epidemic. Classical compartmental models must be modified and studied to correctly describe the effects of forced external actions to reduce the impact of the disease. In addition, data are often incomplete and heterogeneous, so a high degree of uncertainty must naturally be incorporated into the models. In this work we address both these aspects, through an optimal control formulation of the epidemiological model in presence of uncertain data. After the introduction of the optimal control problem, we formulate an instantaneous approximation of the control that allows us to derive new feedback controlled compartmental models capable of describing the epidemic peak reduction. The need for long-term interventions shows that alternative actions based on the social structure of the system can be as effective as the more expensive global strategy. The importance of the timing and intensity of interventions is particularly relevant in the case of uncertain parameters on the actual number of infected people. Simulations related to data from the recent COVID-19 outbreak in Italy are presented and discussed.

## 1 Introduction

From the digital tracking systems of the Koreans to UK’s initial choice of not wanting to do anything to counter the spread of the virus, passing through the militarized quarantines of the Chinese and those less authoritarian and more involved of the Italians, the reaction of different countries to the COVID-19 outbreak has shown a series of very different approaches that can also be explained considering the different cultural and political attitudes of the countries concerned.

In all cases, after an initial phase, even those governments that were less restrictive in the face of the pandemic’s inexorable progress had to take strong containment measures. There’s a graph that has become the symbol of the COVID-19 pandemic most of all. It shows in a simple and intuitive way the importance of slowing down the spread of an epidemic as much as possible (”flattening the curve”), so that the healthcare system can take care of all the sick without collapsing. Its success has helped to save many lives, raising awareness of good practices to slow down an epidemic: stay as much as possible at home, reduce social interactions and wash your hands often and well.

These “non-pharmaceutical” intervention measures, however, entail significant social and economic costs and thus policy makers may not be able to maintain them for more than a short period of time. Therefore, a modelling approach based on a limited time horizon that takes into account the social structure of the population is necessary in order to optimize containment strategies. Most current research, however, has focused on control procedures aimed at optimizing the use of vaccinations and medical treatments [5, 7, 14, 15] and only recently the problem has been tackled from the perspective of non-pharmaceutical interventions [29, 32]. In addition, data collected by governments are often incomplete and heterogeneous, so a high degree of uncertainty needs to be incorporated into predictive models [9, 11, 16, 25, 31, 38]. This is the case of the spreading of COVID-19 worldwide, which have been often mistakenly underestimated due to a combination of factors, including deficiencies in surveillance and diagnostic capacity, and the large number of infectious but asymptomatic individuals [25, 31, 41].

For almost a hundred years, mathematical models have been used to describe the spread of epidemics [27]. The models currently used largely originate from the model proposed by Kermack and McKendrick at the beginning of last century. Even if the model contains strong simplification assumptions, the concepts introduced through this model are essential to provide a first intuition on the dynamics of epidemics, an intuition that remains confirmed in more complex models, albeit with numerous modifications (see for example [10,23]). The model provides for the division of the population into compartments, the susceptible, healthy individuals who may be infected, the infectious, who have already contracted the disease and can transmit it, and the removed, compartment that includes those who are healed and immune.

The hypothesis made by Kermack and Mckendrick is that of the homogeneous “mixing”; that is, it is assumed that each individual has the same probability of contacting any other individual in the population. One understands how this hypothesis is unrealistic: we are often in contact with people from our family, our workplace, school class, group of friends and very rarely with those who live in a different place, have different ages and professions. In recent years, therefore, computational models have been developed that try to take into account additional social characteristics of individuals in order to arrive at more accurate predictions by keeping, however, the simplicity of compartmental models [13,20–22,24,28].

In this paper starting from a general compartmental model with social structure we consider the external action of a policy maker that aims at reducing the spread of the epidemics by applying non pharmaceutical intervention measures, such as social distancing and quarantine. The mathematical problem is formulated as an optimal control problem characterized by a functional cost whose objective is to minimize the number of infectious people in a given time horizon. Through an instantaneous control strategy we compute an explicit feedback control that allows us to derive new SIR-type compartmental models capable of describing the epidemic peak reduction. Previously, this type of approach has been used successfully in the case of social models of consensus [1–4,8,12,18].

The feedback controlled models are subsequently extended to take into account the presence of uncertain infection parameters and data. In fact, to have reliable forecasts it is of paramount importance to consider the presence of uncertain quantities as a structural feature of the epidemic dynamics. From a mathematical point of view, we can rely on the methods of uncertainty quantification (UQ) to obtain efficient and accurate solutions based on stochastic orthogonal polynomials for the differential model with random inputs [40]. Few results are actually available regarding methods of UQ in epidemic systems, we mention in this direction [9, 11, 16, 38]. The main idea is to increase the dimensionality of the problem adding the possible sources of uncertainty from the very beginning of the modelling. Hence, we extrapolate statistics by looking at the so-called quantities of interest, i.e. statistical quantities that can be obtained from the solution and that give some global information with respect to the input parameter like expected solution of the problem or higher order moments. Several techniques can be adopted for the approximation of the quantities of interest, in this paper we adopt stochastic Galerkin methods that allow to reduce the problem to a set of deterministic equations for the numerical evaluation of the solution in presence of uncertainties. We refer the interested reader to recent surveys and monographs on the topic [17,26,34,40].

In particular, we consider the case in which the policy maker applies his control based on several possible estimators on the actual number of infected people. The need for long-term interventions shows that alternative actions based on the social structure of the system can be as effective as the more expensive optimal strategy. The importance of the timing and intensity of interventions is particularly relevant in the case of uncertain parameters on the actual number of infected people.

The rest of the manuscript is organized as follows. In Section 2 we introduce the structured social SIR model and formulate the mathematical approach for containment measures to reduce the spread of the disease. Next, a feedback controlled model used in the subsequent analysis is derived within a short time horizon approximation. In Section 3 we generalize the feedback controlled model to take into account the presence of uncertainties. Section 4 is dedicated to the presentation of some numerical examples including applications to the COVID-19 epidemic in Italy. The most up-to-date information available to date has been used for these simulations. This illustrates the potential application of the model as a real-time forecasting tool. In separate Appendices we provide details of Galerkin’s stochastic method employed to efficiently address the uncertainties and the social interaction matrices characterizing the contact rates.

## 2 Control of epidemic dynamics

The starting model in our discussion is a SIR-type compartmental model in epidemiology with a social structure. The presence of a social structure is in fact essential in deriving appropriate sustainable control techniques from the population for a protracted period, as in the case of the recent COVID-19 epidemic.

### 2.1 Compartmental models with social structure

The heterogeneity of the social structure, which impacts the diffusion of the infective disease, is characterized by the vector 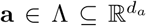 characterizing its social state and whose components summarize, for example, the age of the individual, its number of social connections or its economic status [22,23]. We denote by *s*(**a**, *t*), *i*(**a**, *t*) and *r*(**a**, *t*), the distributions at time *t* > 0 of susceptible, infectious and recovered individuals, respectively in relation to specific social characteristics. We assume that the rapid spread of the disease and the low mortality rate allows to ignore changes in the social structure, such as the aging process, births and deaths.

Consequently, for a given population of total number *N*, we have that

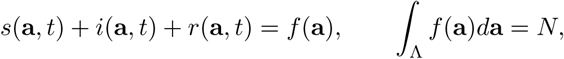

where *f*(**a**) is the total distribution of the social features defined by the vector **a**. Hence, we recover the total fraction of the population which belong to the susceptible, infected and recovered as follows

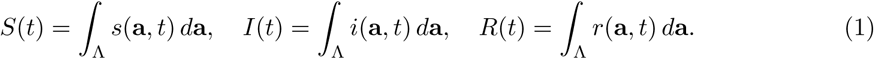

In a situation where changes in the social features act on a slower scale with respect to the spread of the disease, the socially structured compartmental model follows the dynamics

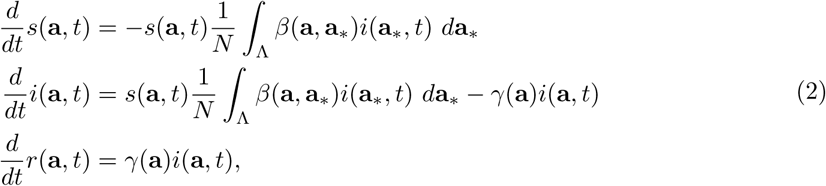

where the function *β*(**a**, **a**_*_) ≥ 0 represents the uncertain interaction rate among individuals with different social features and *γ*(**a**) ≥ 0 the recovery rate which may depend on the social feature.

Often, in socially structured models the interaction rate between people is assumed to be separable, and proportionate to the activity level of the social feature [22, 23], as follows

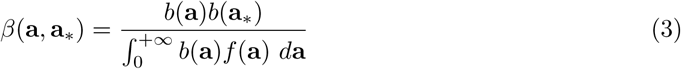

with *b*(**a**) the average number of people contacted by a person with social feature a per unit time. Alternative approaches are based on preferential mixing [13, 21]. Specific examples of age-dependent social interaction matrices are reported in Appendix B.

We introduce the usual normalization scaling

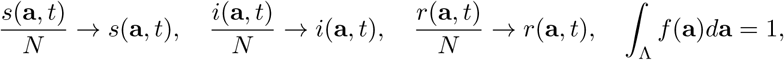

and observe that the quantities *S*(*t*), *I*(*t*) and *R*(*t*) satisfy the conventional SIR dynamic

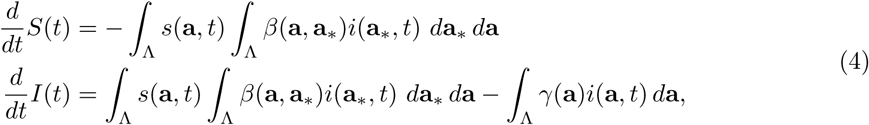

where the fraction of recovered is obtained from *R*(*t*) = 1 − *S*(*t*) − *I*(*t*). We refer to [22, 23] for analytical results concerning model (2) and (4). In the following we will adopt the simple compartmental model (2) to derive our feedback controlled formulation in presence of uncertainty. The extension to more realistic compartmental models in epidemiology such as SEIR and/or MSEIR can be carried out in a similar way.

In order to simplify the description, we will consider the case *d_a_* = 1 and set the social dependence as the age *a* of the individual because of its importance in epidemic dynamics. It is clear, however, that similar containment procedures can be applied also on the basis of other social features. We will first formulate the feedback controlled SIR model in the deterministic case and subsequently extend our approach to the presence of uncertain parameters.

### 2.2 Optimal control of structured compartmental model

In order to define the action of a policy maker introducing a control over the system based on social distancing and other containment measures linked to the social structure we consider an optimal control framework. The choice of an appropriate functional is problem dependent [15].

In our setting, we account the minimization of the total number of the infected population *I*(*t*) through the an age dependent control action depending both on time and pairwise interactions among individuals with different ages.

Thus, we introduce the optimal control problem

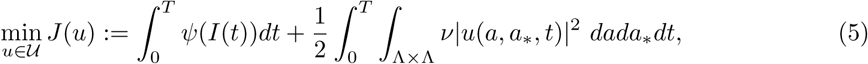

subject to

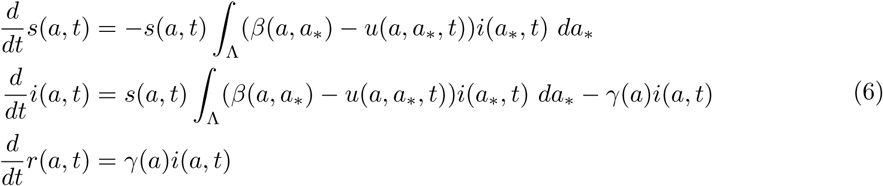

with initial condition *i*(*a*, 0) = *i*_0_(*a*), *s*(*a*, 0) = *s*_0_(*a*) and *r*(*a*, 0) = *r*_0_(*a*).

The number of infected individual is measured by a monotone increasing function *ψ*(·) such that 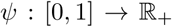. This function models the policy maker’s perception of the impact of the epidemic by the number of people currently infected. For example *ψ*(*I*) = *I^q^/q*, for *q*> 1 implies an underestimation of the actual number of infected that we expect to result in the need for a larger penalty term than *q* = 1. The control aims to minimize this measure of the total infected population by reducing the rate of interaction between individuals. We consider a quadratic cost for its actuation.

Such control is restricted to the space of admissible controls

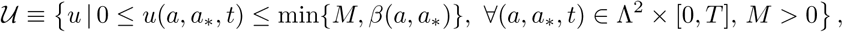

which ensure the admissibility of the solution for (6).

The solution to problem (5)-(6) is computed through the optimality conditions obtained from the Euler-Lagrangian as follows

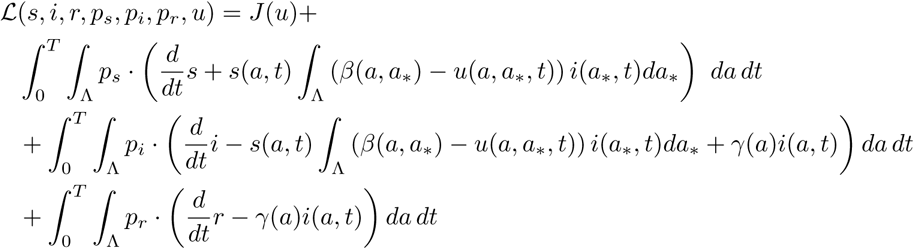

where *p_s_*(*a, t*), *p_i_*(*a, t*), *p_r_*(*a, t*) are the associated lagrangian multipliers. Computing the variations with respect to (*s, i, r*) we retrieve the adjoint system

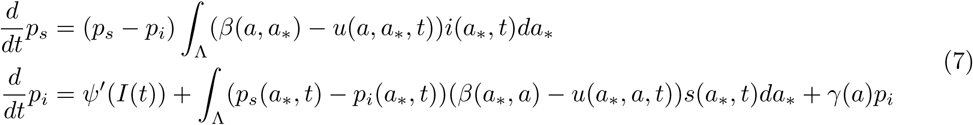

with terminal conditions *p_s_*(*a, T*) = 0, *p_i_*(*a, T*) = 0 and *p_r_*(*a, T*) = 0. Note that the contribution of *p_r_*(*a, t*) vanishes since the control does not act directly on population *R*, and the removed population is not considered in the minimization of the functional. The optimality condition reads

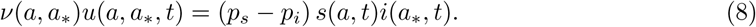

The optimality conditions (7)-(8) are first order necessary conditions for the optimal control *u^*^*(*a, t*). In order to be admissible then the control reads

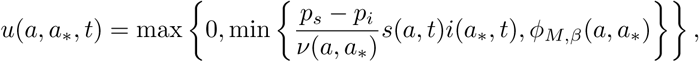

where *ϕ_M,β_*(*a, a_*_*) = min{*β*(*a, a_*_*),*M*}.

The approach just described, however, is generally quite complicated when there are uncertainties as it involves solving simultaneously the forward problem (5)-(6) and the backward problem (7)-(8). Moreover, the assumption that the policy maker follows an optimal strategy over a long time horizon seems rather unrealistic in the case of a rapidly spreading disease such as the COVID-19 epidemic.

### 2.3 Feedback controlled compartmental models

In this section we consider short time horizon strategies which permits to derive suitable feedback controlled models. These strategies are suboptimal with respect the original problem (5)-(6) but they have proved to be very successful in several social modeling problems [1–4, 18]. To this aim, we consider a short time horizon of length *h*> 0 and formulate a time discretize optimal control problem through the functional *J_h_*(*u*) restricted to the interval [*t, t* + *h*], as follows

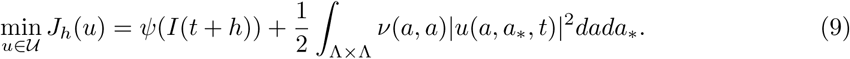

subject to

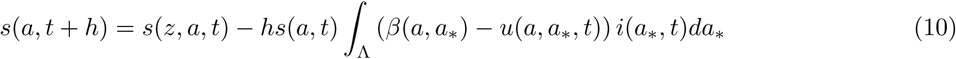

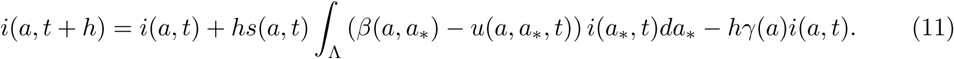

Recalling that the macroscopic information on the infected is

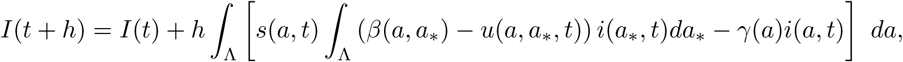

we can derive the minimizer of *J_h_* computing *D_u_J*(*u*) ≡ 0. We retrieve the following nonlinear equation

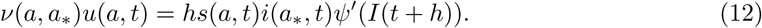

Introducing the scaling *ν*(*a, a_*_*) = *hκ*(*a, a_*_*) we can pass to the limit for *h* → 0. Hence (12) reduces to the instantaneous control

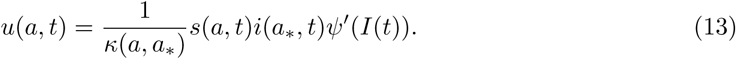

The resulting controlled dynamics is retrieved by applying the instantaneous strategy directly in the continuous system (6) as follows

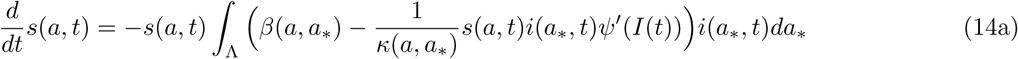

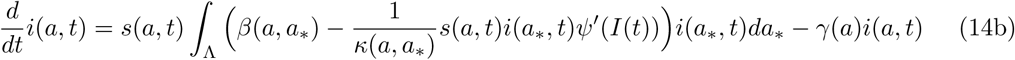

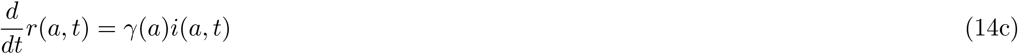

In what follows we provide a sufficient conditions for the admissibility of the instantaneous control in terms of the penalization term *κ*(*a, a_*_*). Indeed we want to assure that the dynamics preserve the monotonicity of the number of susceptible population *s*(*a, t*).

#### Proposition 2.1

*Let β*(*a*, *a_*_*) ≥ *δ* > 0, *then for all* (*a*, *a*_*_) ∈ Λ×Λ *solutions to* (14) *are admissible if the penalization κ satisfies the following inequality*

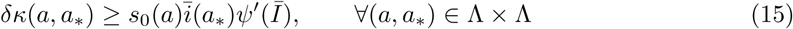

where 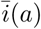 and 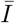 are respectively the peak reached by the infected of class a and by the total population.

*Proof:* By imposing the non-negativity of the controlled reproduction rate inside the integral we have

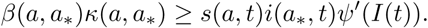

This inequality has to be satisfied for every time *t* ≥ 0. Second we observe that the number of susceptible *s*(*a, t*) is decreasing in time therefore *s*_0_(*a*) ≥ *s*(*a, t*) for all *t*. Moreover *i*(*a, t*) reaches a peak before decreasing to 0 (note that this peak can also be in *t* = 0), say 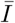 for the macroscopic variable and 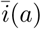. Thus, thanks to the monotonicity of *ψ*(·), we can restrict the previous inequality as follows

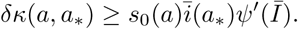

In Figure 1 we report the phase diagram of susceptible-infected trajectories for the controlled model with homogeneous mixing

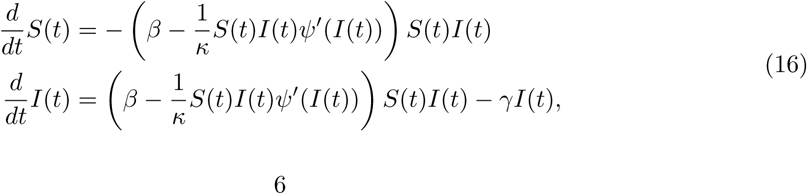

with *ψ*(*I*) = *I^q^/q*. The dynamic is similar to the classical SIR model but with a nonlinear contact rate. In particular, the trajectories are flattened when the value of the control is such that *κβ* ≈ *S*(*t*)*I*(*t*)*ψ′*(*I*(*t*)) and the reproductive number is close to zero. Note, however that this status is not an equilibrium point of the system.

**Figure 1:**
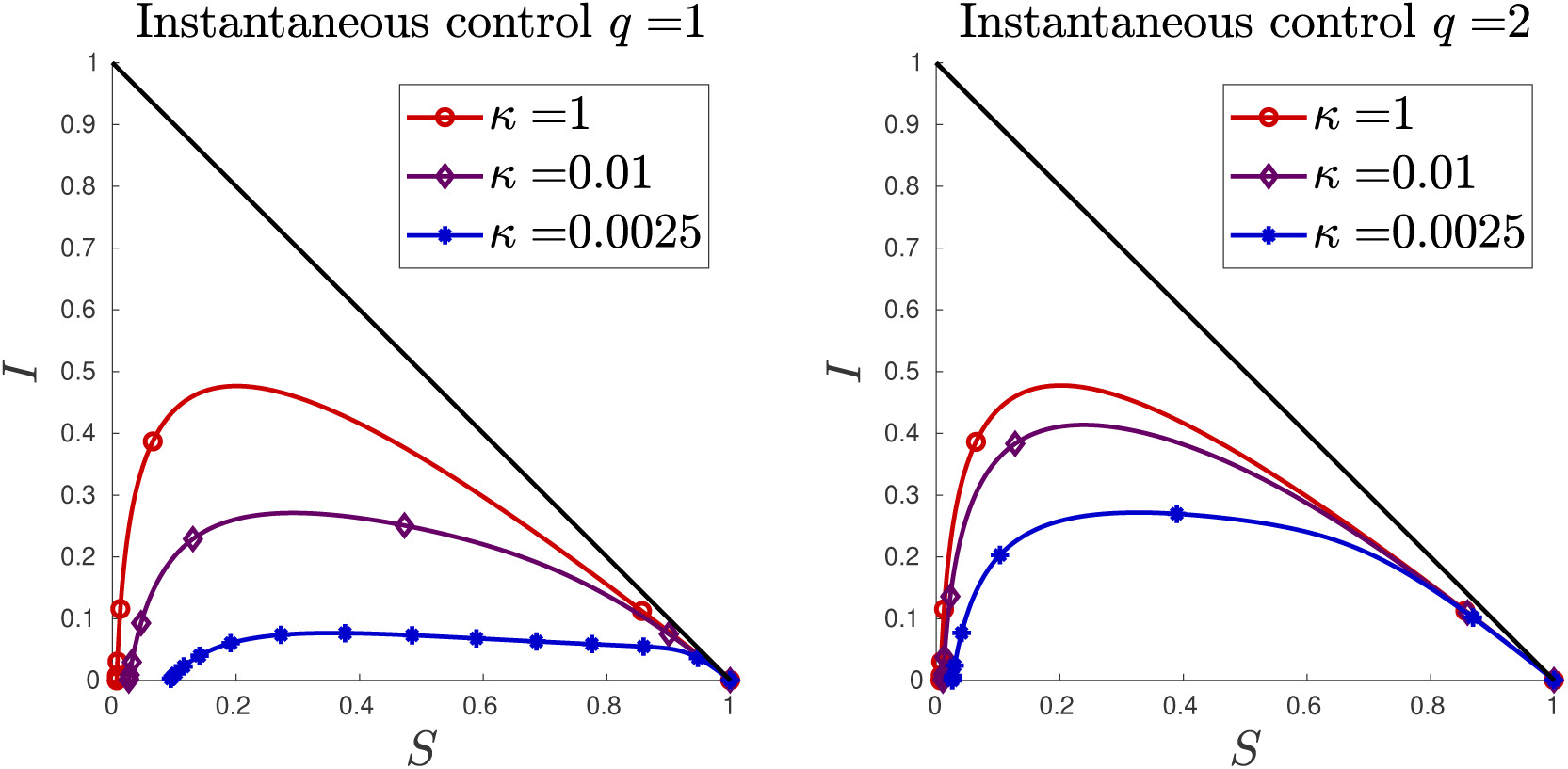
Phase diagram of susceptible-infected trajectories for the controlled SIR-type model with homogenous mixing and *ψ*(*I*) = *I^q^/q*. Different choices of the penalization term *κ* are reported. Left plot the case *q* = 1, right plot *q* = 2.

To understand this, let us observe that an equilibrium state (*S*^*^, *I*^*^) for (16) satisfies the equations

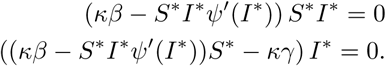

An equilibrium point corresponds to the classical state in which we have the extinction of the disease *I*^*^ = 0 and *S*^*^ arbitrary and defined by the asymptotic state of the dynamics [23]. Now, let’s suppose that *I*^*^ ≠, *S*^*^≠ 0, we can look for solutions where control is able to perfectly balance the spread of the disease

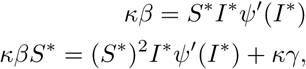

consequently

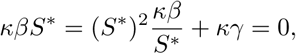

which is satisfied only for *γ* = 0 when *κ* ≠ 0.

## 3 Control of epidemic dynamics with uncertainties

Since the beginning of the outbreak of new infectious diseases, the actual number of infected and recovered people is typically underestimated, causing fatal delays in the implementation of public health policies facing the propagation of epidemic fronts. This is the case of the spreading of COVID-19 worldwide, often mistakenly underestimated due to deficiencies in surveillance and diagnostic capacity [25,37]. Health systems are struggling to adopt systematic testing to monitor actual cases. Moreover, another important epidemiological factor that pollutes the available data is the proportion of asymptomatic [25, 31, 41].

Among the common sources of uncertainties for dynamical systems modeling epidemic outbreaks we may consider the following

- noisy and incomplete available data
- structural uncertainty due to the possible inadequacy of the mathematical model used to describe the phenomena under consideration.

In the following we consider the effects on the dynamics of uncertain data, such as the initial conditions on the number of infected people or the interaction and recovery rates. On the numerical level we consider techniques based on stochastic Galerkin methods, for which spectral convergence on random variables is obtained under appropriate regularity assumptions [40]. For simplicity, the details of the numerical method that allows to reduce the uncertain dynamic system to a set of deterministic equations are reported in the Appendix A.

### 3.1 Socially structured models with uncertain inputs

We introduce the random vector 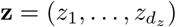 whose components are assumed to be independent real valued random variables

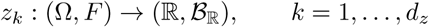

with 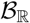 the Borel set. We assume to know the probability density 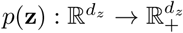 characterizing the distribution of **z**. Here, 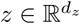 is a random vector taking into account various possible sources of uncertainty in the model.

In presence of uncertainties we extend the initial modelling by introducing the quantities *s*(**z**, *a, t*), *i*(**z**, *a, t*) and *r*(**z**, *a, t*) representing the distributions at time *t* ≥ 0 of susceptible, infectious and recovered individuals. The total size of the population is a deterministic conserved quantity in time, i.e.

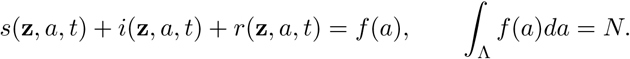

Hence, the quantities

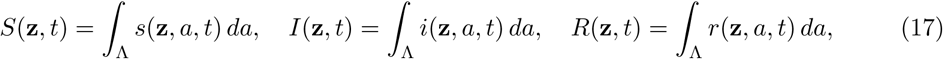

denote the uncertain fractions of the population that are susceptible, infectious and recovered respectively.

The social structured model with uncertainties reads

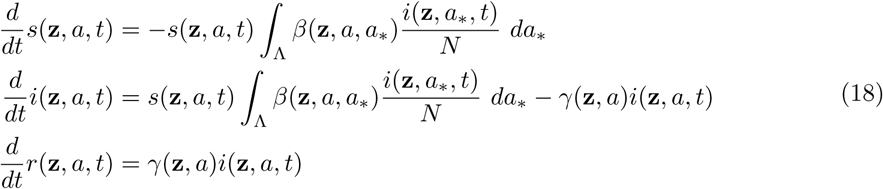

To illustrate the impact of uncertainties let us consider the simple following example. In the case of homogeneous mixing with uncertain contact rate *β*(*z*) = *β* + *αz*, *α*> 0, *z* ∈ ℝ distributed as *p*(*z*) the model reads

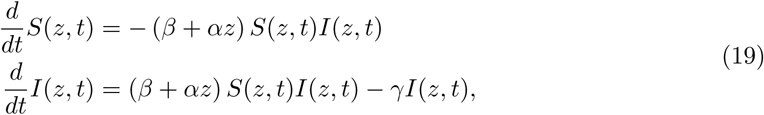

with deterministic initial values *I*(*z*, 0) = *I*_0_ and *S*(*z*, 0) = *S*_0_. The solution for the proportion of infectious during the initial exponential phase is [38]

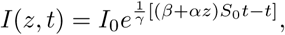

and its expectation

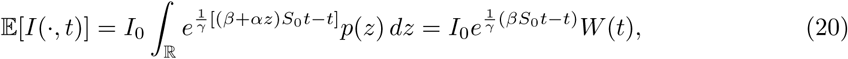

where the function

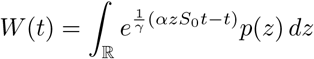

represents the statistical correction factor to the standard deterministic exponential phase of the disease 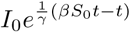. If *z* is uniformly distributed in [−1, 1] we can explicitly compute

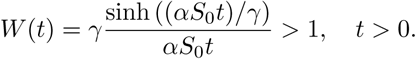

More in general, if *z* has zero mean then by Jensen’s inequality we have *W* (*t*) > 1 for *t*> 0, so that the expected exponential phase is amplified by the uncertainty (see [38]).

In a similar way, keeping *β* constant, but introducing a source of uncertainty in the initial data *I*(*z*, 0) = *I*_0_ + *µz*, *µ*> 0 and *z* ∈ ℝ distributed as *p*(*z*) the solution in the exponential phase reads

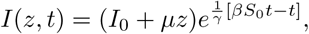

and then its expectation

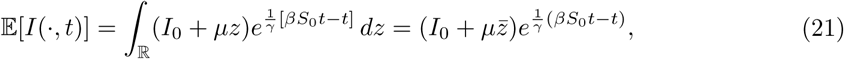

where 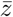 is the mean of the variable *z*. Therefore, the expected initial exponential growth behaves as the one with deterministic initial data 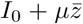. Of course, if both sources of uncertainty are present the two effects just described sum up in the dynamics.

### 3.2 The feedback controlled model with random inputs

In presence of uncertainties the optimal control problem (5)-(6) is modified as follows

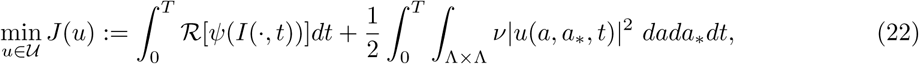

being 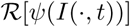 a suitable operator taking into account the presence of the uncertainties **z**. Examples of such operator that are of interest in epidemic modelling are the expectation with respect to uncertainties

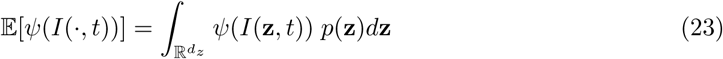

or relying on data which underestimate the number of infected

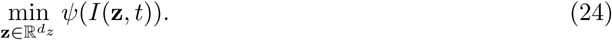

In (22) 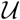 the space of admissible controls is defined as

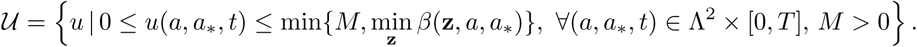

The above minimization is subject to the following dynamics

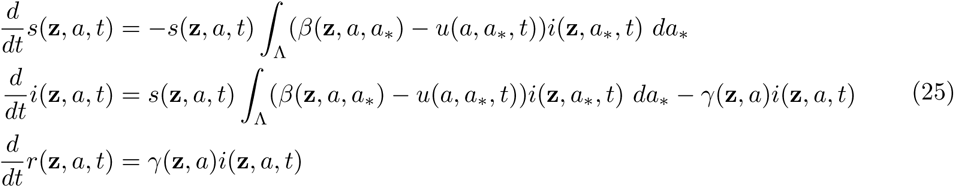

with initial condition *i*(**z**, *a*, 0) = *i*_0_(**z**, *a*), *s*(**z**, *a*, 0) = *s*_0_(**z**, *a*) and *r*(**z**, *a*, 0) = *r*_0_(**z**, *a*).

The implementation of instantaneous control for dynamics in presence of uncertainties follows from the derivation presented in Section 2.3 and we omit the details. The resulting feedback control *u*(*a, a*_*_, *t*) reads

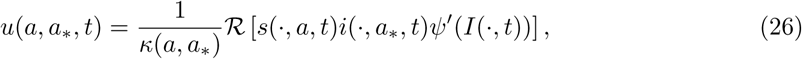

and defines the feedback controlled model in presence of uncertainties.

## 4 Examples from the COVID-19 outbreak in Italy

In this section we present several numerical tests on the constrained compartmental model with uncertain data. Details of the numerical method used are given in Appendix A. In an attempt to analyse sufficiently realistic scenarios, in the following examples we will refer to values taken from Italian data on the COVID-19 epidemic [36]. More precisely, in the first test case we illustrate the behavior of the model in a simplified approach in the absence of uncertainty and social structure and without trying to reproduce scenarios closely related to current data. In the second test case, following a progressively more realistic approach, we consider the impact of the presence of uncertain data in the controlled model with homogeneous social mixing and calibrated on Italian data. The same setting is then considered in Test 3 taking into account the additional effect on the spreading of the infectious disease given by the social structure of the system described by suitable social interaction functions. The final scenario, explored in Test 4, examines the possibility of reducing the amplitude of the epidemic peak with the application of relaxed confinement measures related to the social structure of the system.

### 4.1 Test 1. Containment in homogeneous social mixing dynamics

To illustrate the effects of controls introduced that mimic containment procedures, let us first consider the case where the social structure is not present. Furthermore, to simplify further the modeling, in this first example we neglect any dependence on uncertain data.

We consider as initial small number of infected and recovered *i*(0) = 3.68 × 10^−6^, *r*(0) = 8.33 × 10^−8^. These normalized fractions refer specifically to the first reported values in the case of the Italian outbreak of COVID-19, even if in this simple test case we will not try to match the data in a predictive setting but simply to illustrate the behavior of the feedback controlled model.

Based on recent studies [25, 41], the initial infection rate of COVID-19 has been estimated of an *R*_0_ = *β/γ* between 2 and 6.5. Here, to exemplify the possible evolution of the pandemic we consider a value close to the lower bound, taking *β* = 25 and *γ* = 10, so that *R*_0_ = 2.5.

In Figures 2 and 3 we report the infected and recovered dynamics based on the activation of the control in two different time frames. In Figure 2 the activation for *t* ∈ [0.5, 1], which means that for *t* > 1 we suppose that the containment restrictions are deactivated. In Figure 3 we consider a larger activation time frame *t* ∈ [0.5, 2].

**Figure 2:**
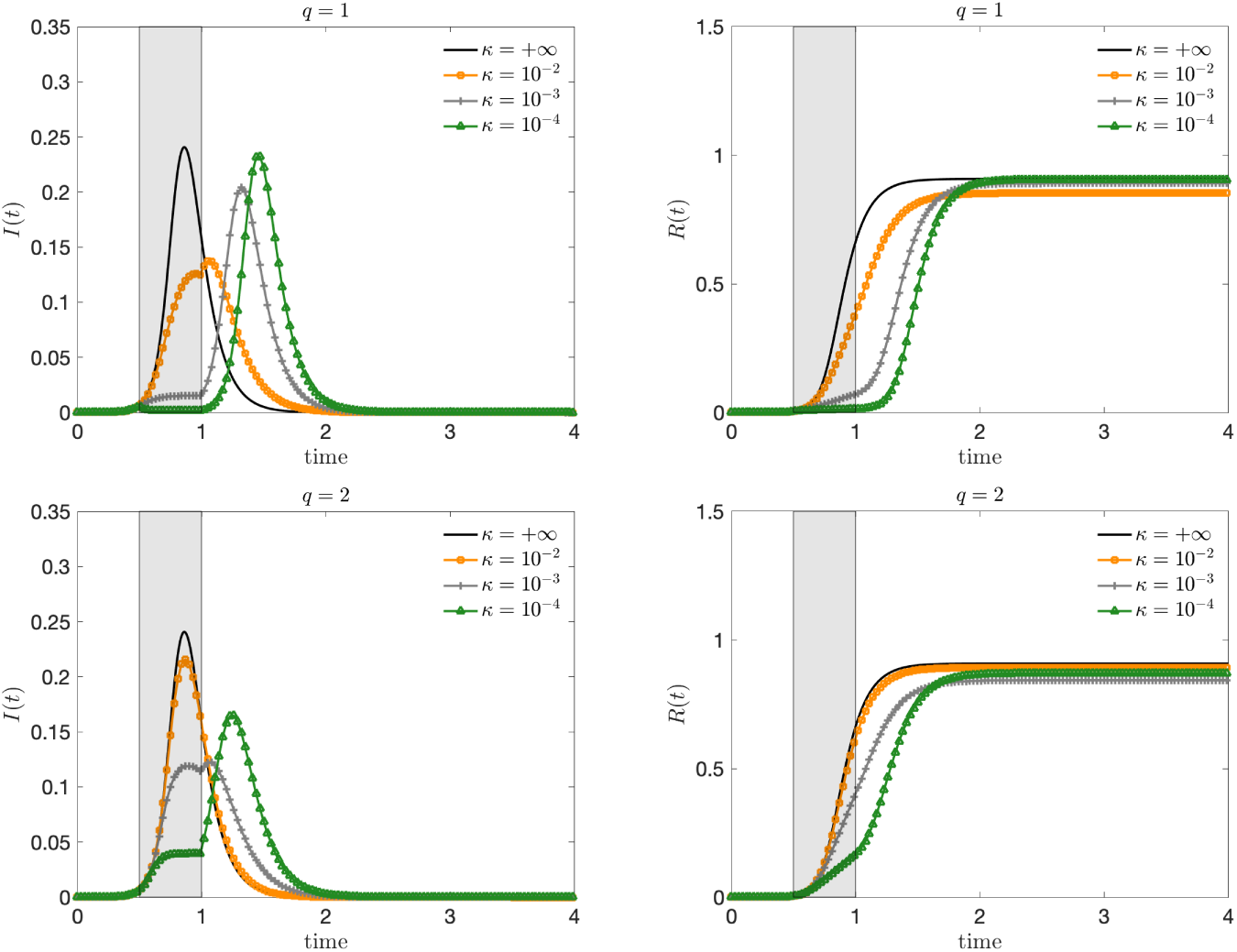
**Test 1**. Evolution of the fraction of infected (left) and recovered (right) based on the constrained model in the interval *t* ∈ [0.5, 1] for *ψ*(*I*) = *I^q^/q*, *q* = 1, 2 and several penalization of the control *κ* = 10^−2^, 10^−3^, 10^−4^. The choice *κ* =+∞ corresponds to the unconstrained case.

**Figure 3:**
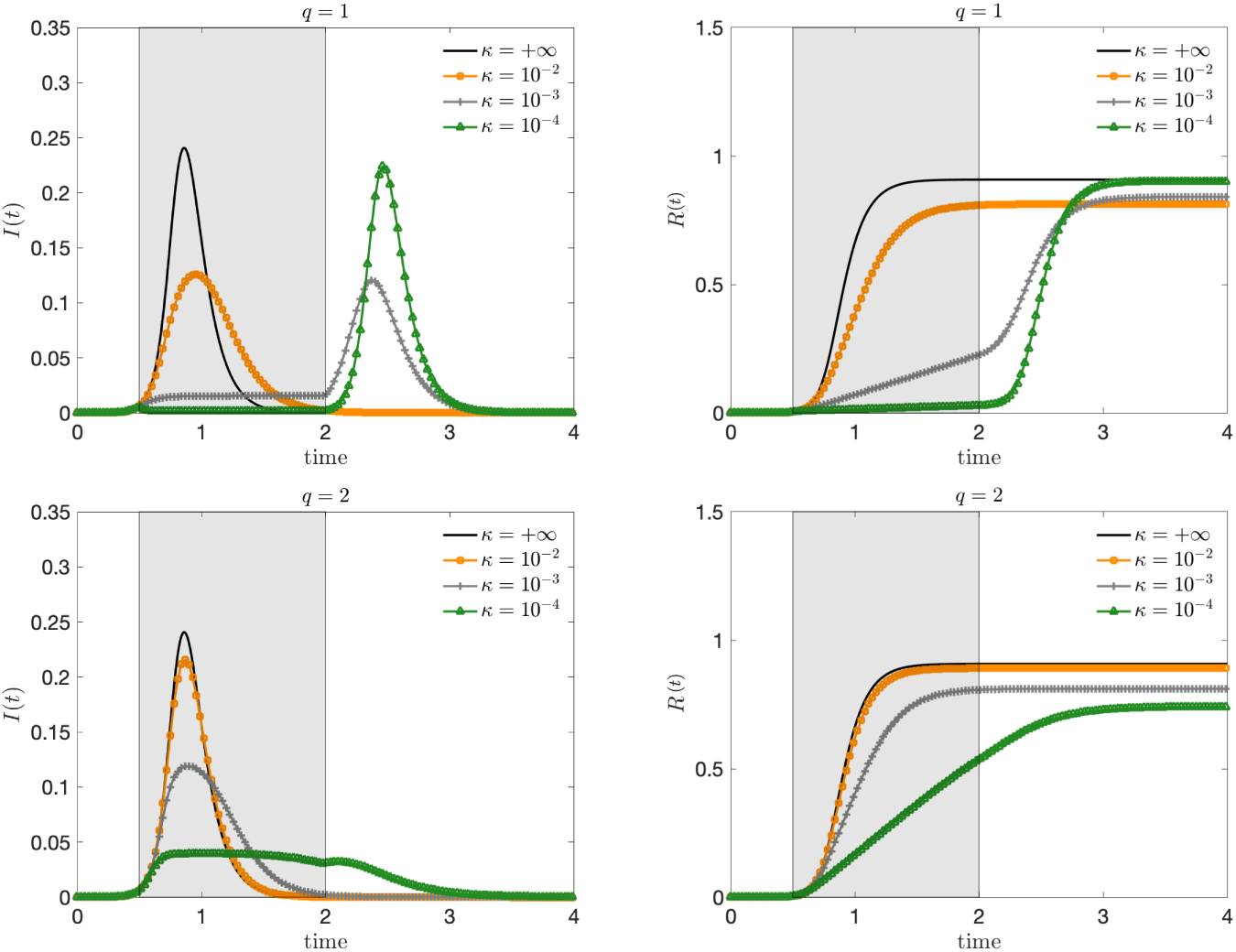
**Test 1**. Evolution of the fraction of infected (left) and recovered (right) based on the constrained model in the interval *t* ∈ [0.5, 2] for *ψ*(*I*) = *I^q^/q*, *q* = 1, 2 and several penalization of the control *κ* = 10^−2^, 10^−3^, 10^−4^. The choice *κ* =+∞ corresponds to the unconstrained case.

With the choice of *ψ*(*I*) = *I^q^/q*, *q* ≥ 1, we can observe how the control term is able to flatten the curve even if, as expected, the case *q* = 2 gives rise to a weaker control action. Note, however, that if the activation time is too short the control is not able to significantly change the total number of infected (and therefore recovered). On the other hand, by enlarging the activation time in combination with a sufficiently small penalty constant, the peak infection is not only reduced, but the total number of infected people is decreased. To achieve this, the control should be kept activated for a sufficiently long time and with the right intensity in a kind of plateau regime where there is a perfect balance between the containment effect and the spread of the disease. On the contrary, if the control is too strong, the majority of the population remains susceptible and consequently the disease will start spreading again forming a second wave after the containment policy is removed.

The cost functional depends on the value of *q* and can be evaluated summing up contributions in (9) with explicit form of the control given by (13). In Figure 4 the cost of the two interventions is compared. We can see how a higher cost is associated with *q* = 1 that can be obtained with the control *q* = 2 for weaker penalizations. Then, in Figure 5 we compare the performance of the two controls with a deactivation time in the range *t* ∈ [1, 3]. To align the costs of interventions, we consider *κ* = 10^−3^ for *q* = 1 and *κ* = 10^−4^ for *q* = 2. It can be observed that there is a minimum control horizon for both strategies, in order to avoid the onset of a second infection peak. A sufficiently long control horizon is therefore necessary to reduce the impact of the infection.

**Figure 4:**
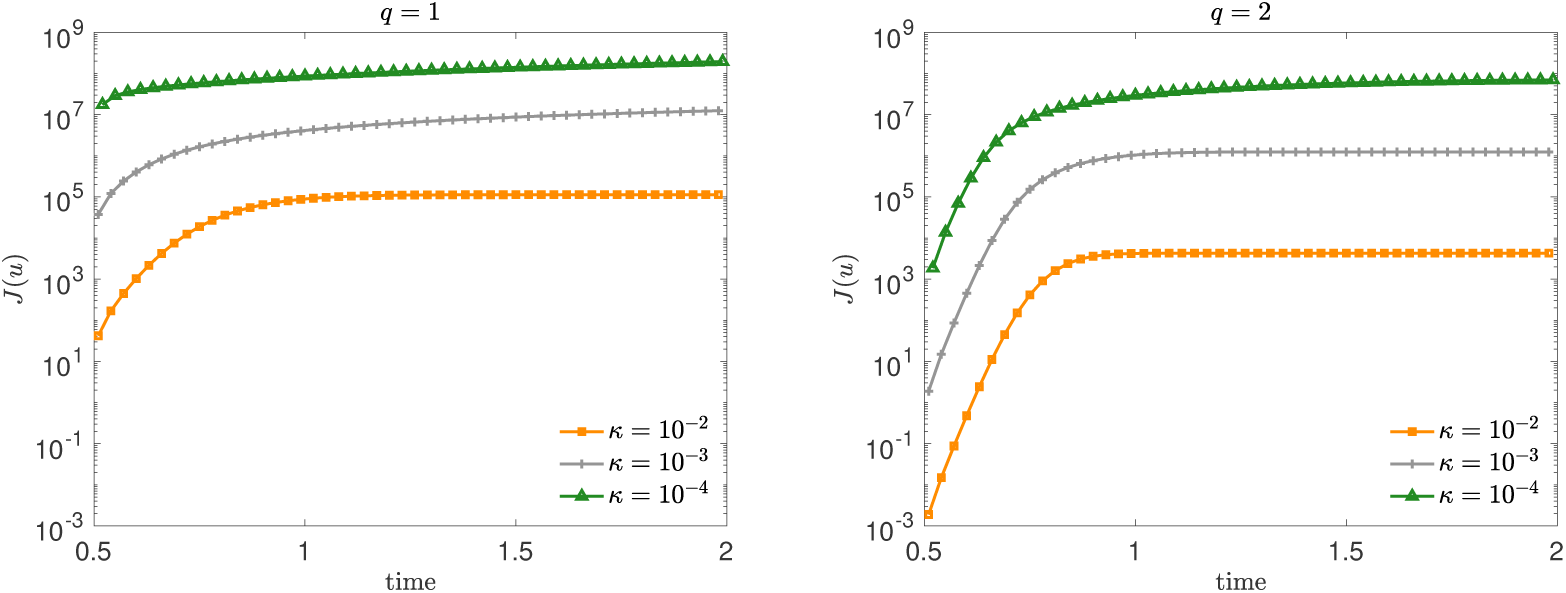
**Test 1**. Evaluation of the costs functional *J*(*u*) for *ψ*(*I*) = *I^q^/q*, *q* = 1 (left) and *q* = 2 (right) on the activation frame *t* ∈ [0.5, 2] for the dynamic in Figure 3. We considered three penalizations *κ* = 10^−2^, 10^−3^, 10^−4^.

**Figure 5:**
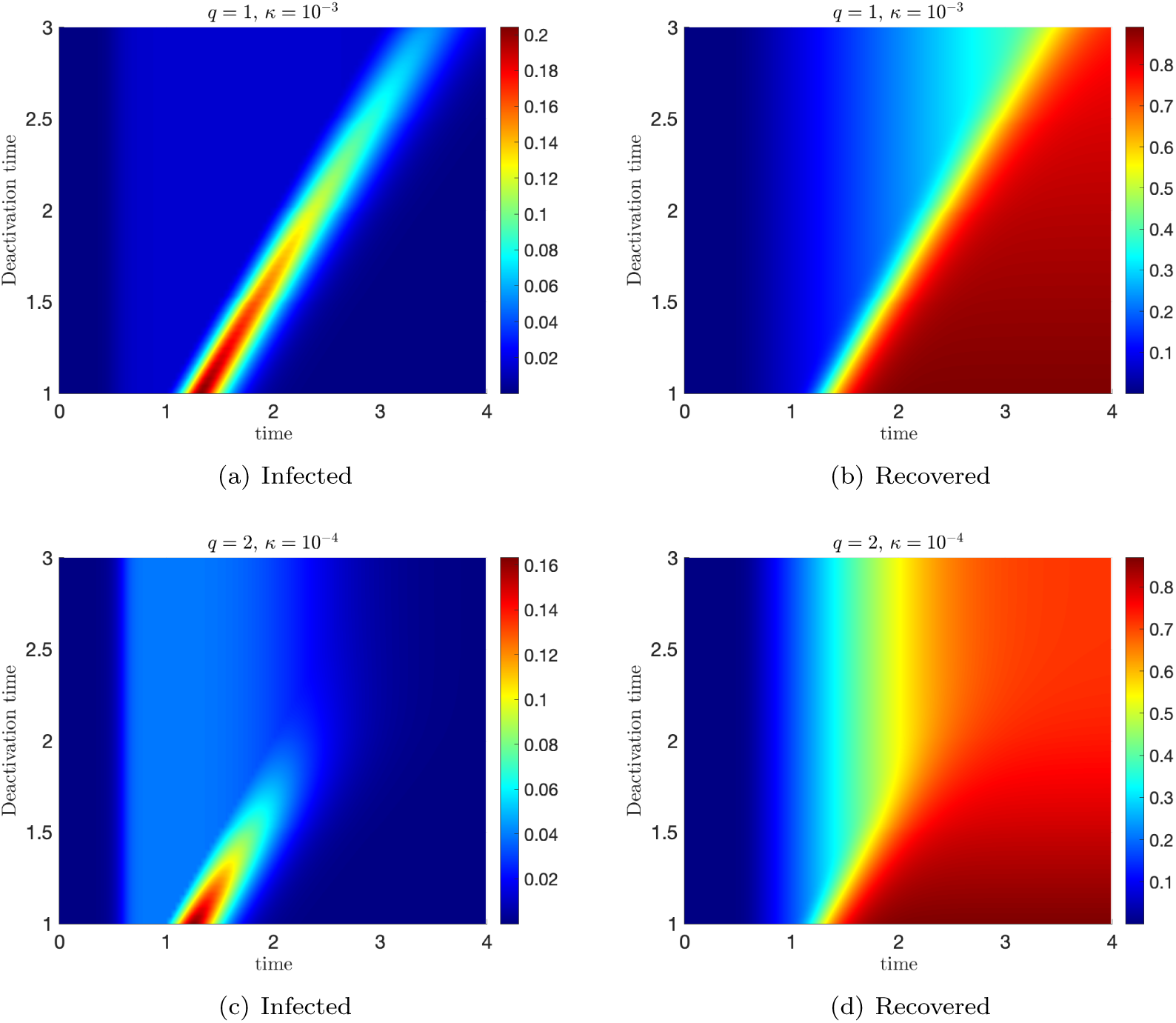
**Test 1**. Evolution of the constrained model for *ψ*(*I*) = *I^q^/q* and *q* = 1 (left column) and *q* = 2 (right column) for a deactivation time of the control in [1, 3]. We considered two penalizations *κ* = 10^−3^ for *q* = 1 and *κ* = 10^−4^ for *q* = 2 in order to align the costs of interventions.

### 4.2 Test 2: Impact of uncertain data on the epidemic outbreak

Next we focus on the influence of uncertain quantities on the controlled system with homogeneous mixing focusing on available data for COVID-19 outbreak in Italy, see [36]. According to recent results on the diffusion of COVID-19 in many countries the number of infected, and therefore recovered, is largely underestimated on the official reports, see e.g. [25, 31].

The estimation of epidemiological parameters is a very difficult problem that can be addressed with many different approaches [9, 16, 38]. In our case, we have limited ourselves to identifying the deterministic parameters of the model through a standard fitting procedure, considering the possible uncertainties due to such an estimation as part of the subsequent uncertainty quantification process. It has been reported, in fact, that deterministic methods based on compartmental models overestimate the effective reproduction number [30].

In order to calibrate the models with the reported quantities we solved two separate constrained optimization problems in absence of uncertainties. First we estimated *β* and *γ* by considering on the time interval [*t*_0_, *t_u_*] the following least square problem

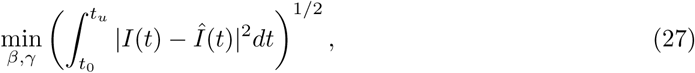

namely the *L*^2^ norm of the difference between the observed number of infected 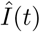 and the theoretical evolution of the unconstrained model (*κ* =+∞). Problem (27) has to be solved under the constraints *β* ≥ 0, *γ* ≥ 0. This problem is solved over the time span [*t*_0_, *t_u_*] where we assumed no social containment procedure was activated.

Next, we estimate the penalization *κ* = *κ*(*t*) in time by solving over a sequence of time steps *t_i_* of size *h* in the controlled time interval *t* ∈ [*t_u_,t_c_*] the problem

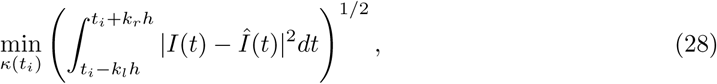

under the constraint *κ*(*t_i_*) > 0, *k_l_,k_r_* ≥ 1 integers, and where for the evolution we consider the values *β* and *γ* estimated in the first optimization step (27). In details, since the available data start on February 24 2020, when no social restrictions were enforced by the Italian government, and since the lockdown started on March 9 we considered *t_u_* = 14 (days). The second fitting procedure has been activate up to last available data with daily time stepping (*h* = 1) and a window of eight days (*k_l_* = 3, *k_r_* = 5) for regularization along one week of available data. To solve problem (27) we tested different optimization methods in combination with adaptive solvers for the system of ODEs. Once calibrated the model we estimated *β* ≈ *β_e_* = 31.2 and *γ* ≈ *γ_e_* = 4.99 which corresponds to an initial attack rate *R*_0_ ≈ 6.25 which agrees with other observations [30]. The corresponding time dependent values for the control in the case *ψ*(*I*) = *I^q^/q* are reported in Figure 6. After an initial adjustment phase the penalty terms converge towards a constant value that we can assume as fixed in predictive terms for future times in a lockdown scenario. This is consistent with a situation in which society needs a certain period of time to adapt to the lockdown policy.

In order to have an insight on global impact of uncertain parameters we consider a two-dimensional uncertainty **z** = (*z*_1_, *z*_2_) with independent components such that

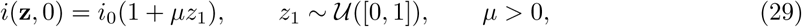

and

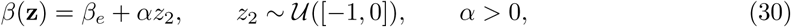

where *i*_0_ is the same as in Test 1 and taken from [36] on February 24, 2020 and *β_e_ an*d *γ_e_ ar*e estimated from (27). Of course, other probability distributions *p*(**z**) = *p*_1_(*z*_1_)*p*_2_(*z*_2_) defined only for *z*_1_, *z*_2_ ≥ 0 are possible, like beta distributions [38, 40]. However, qualitatively the results do not change and the introduction of a beta distribution would imply dependence on an additional parameter for each random variable, so we limit ourselves to the simple situation of uniform uncertainty.

**Figure 6:**
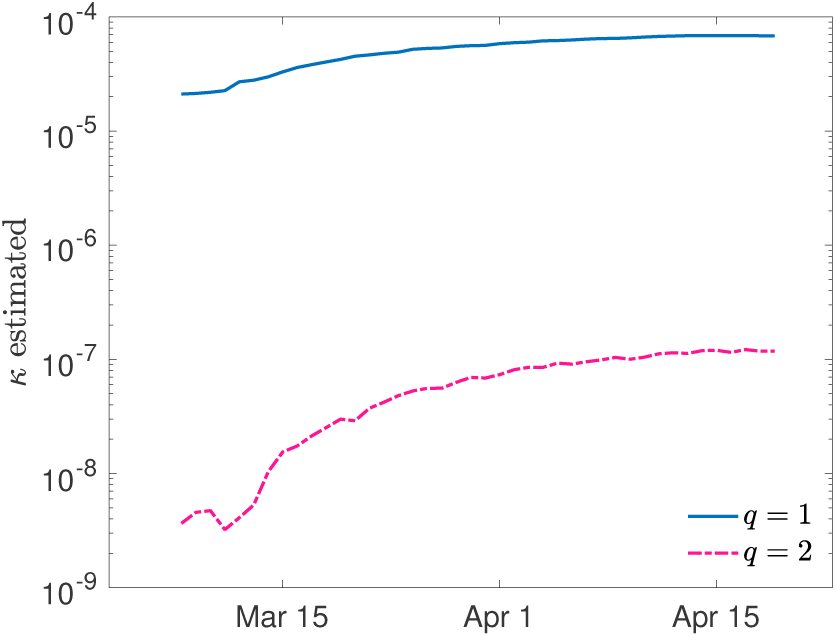
**Test 2**. Estimated control penalization terms over time from reported data on number of infected people in the case of COVID-19 outbreak in Italy.

In all the considered evolutions we adopted a stochastic Galerkin approach, see Appendix A with *M* = 10 and a fourth order Runge-Kutta method for the time integration. The feedback controlled model has been computed using as estimation of the total number of infected reported, namely

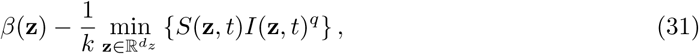

in agreement with the fitting procedure obtained from the lower bounds of the uncertain initial data.

In Figure 7 we represent the evolution of the expected value of the number of infected obtained by the controlled model in the presence of initial random data (29) and uncertain contact frequency (30). The value *µ* = 10 have been chosen accordingly to the WHO suggestions that around 80% are asymptomatic ^1^. The uncertainty in the contact rate has been modeled taking *α* = 1.

**Figure 7:**
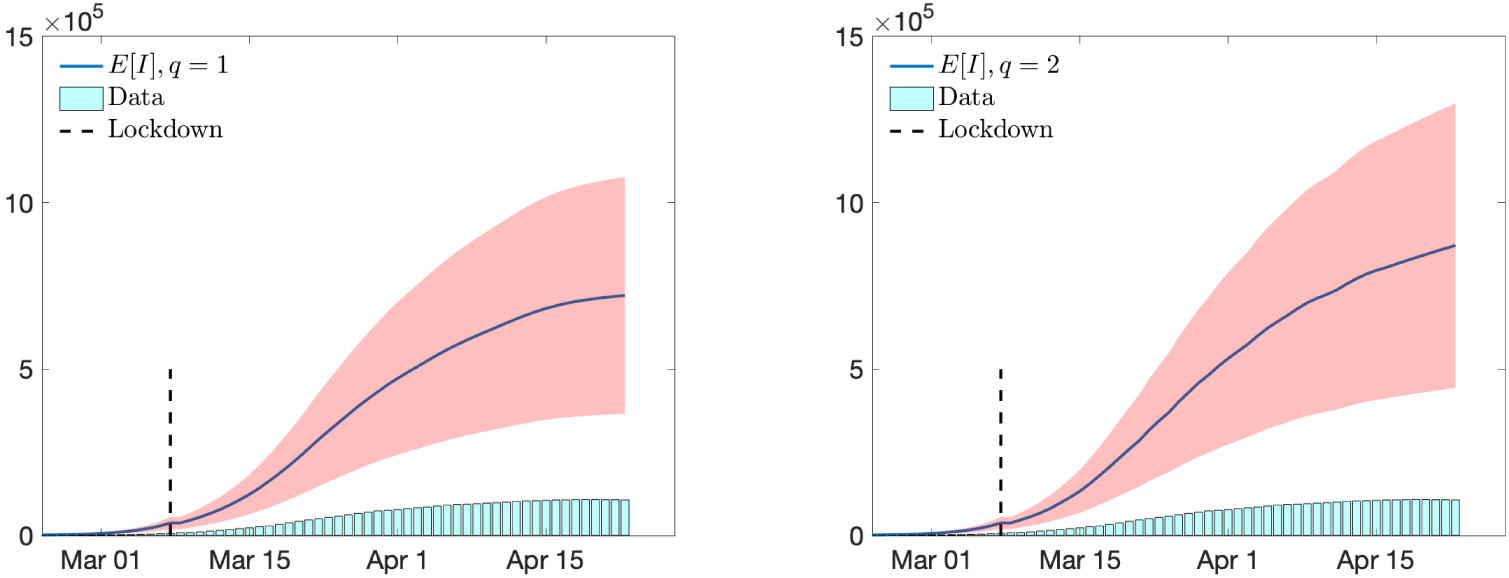
**Test 2**. Evolution of expected number of infected and their confidence bands for the calibrated control model with *ψ*(*I*) = *I^q^/q*, *q* = 1, 2 with uncertain initial data (29) with *µ* = 10 and uncertain reproduction number (30) with *α* = 1 in the case of COVID-19 outbreak in Italy.

We represented the expected values of the number of infected along with the confidence bands obtained from the overall variance in **z**. The range of true infected as seen increases dramatically in both cases *q* = 1 and *q* = 2 with a strong uncertainty on the effective numbers. The bars below the graph are the reported values of the number of infected on which the model has been calibrated.

From (31) we can estimate the effect of the lockdown policies on the value of *R*_0_ in time. The effect of the uncertain initial estimated value *R*_0_ is translated in the confidence bands from *z*_2_ reported in the left plot of Figure 8. The results show that the *R*_0_ reproduction number, thanks to drastic containment actions, has been drastically reduced and its expected value is now just below one. This shows the importance to continue with containment policies to avoid a restart of the spread of the epidemic.

**Figure 8:**
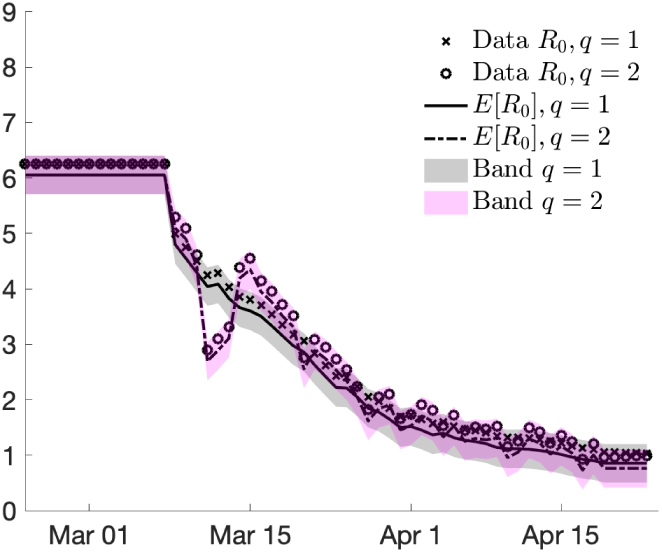
**Test 2**. Estimated reproduction number *R*_0_ from the controlled model for *ψ*(*I*) = *I^q^*, *q* = 1 and *q* = 2 together with the confidence bands in the case of COVID-19 outbreak in Italy.

### 4.3 Test 3: The effect of social contacts in the population

We analyze the effects of the inclusion of age dependence and social interactions in the above scenario. More precisely we consider the social interaction functions reported in Appendix B and uncertain initial number of infected. These functions were normalized using the previously estimated parameters *β_e_* and *γ_e_* in accordance with

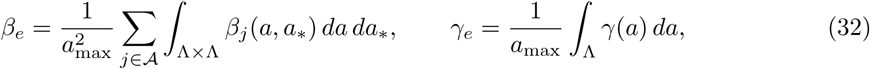

where Λ = [0, *a*_max_], *a*_max_ = 100 and where *γ*(*a*) accordingly to recent studies on age-related recovery rates [39] has been chosen as a decreasing function of the age as follows

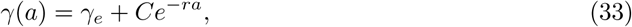

with *r* = 5 and *C* ∈ ℝ such that (32) holds. Clearly, this choice involves a certain degree of arbitrariness since there are not yet sufficient studies on the subject, nevertheless, as we will see in the simulations, it is able to reproduce more realistic scenarios in terms of age distribution of the infected without altering the behaviour relative to the total number of infected.

We divided the computation time frame into two zones and used different models in each zone, in accordance with the policy adopted by the Italian Government. The first time interval defines the period without any form of containment from 24 February to 9 March, the second the lockdown period from 9 March. In the first zone we adopted the uncontrolled model with homogeneous mixing for the estimation of epidemiological parameters. Hence, in the second zone we compute the evolution of the feedback controlled age dependent model (25)-(26) with matching (on average) interaction and recovery rates (32) and with the estimated control penalization *κ*(*t*) as reported in Figure 6 until April 23rd. After April 23rd the computation advances in time using as penalization term the constant asymptotic value 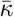 reached by *k*(*t*). The initial values for the age distributions of susceptible and infectious individuals are shown in Figure 9 in agreement with reported data^2^.

**Figure 9:**
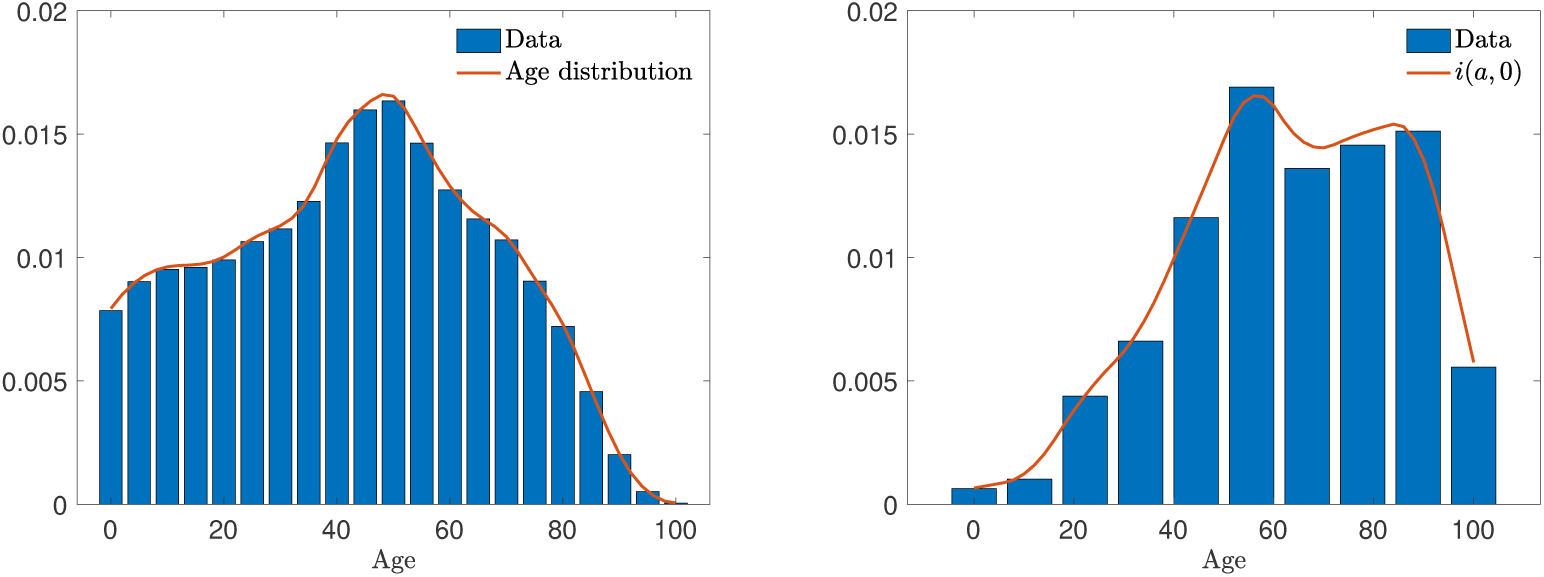
**Test 3**. Distribution of age in Italy (left) and distribution of infected (right) together with the corresponding continuous approximations^2^.

In Figure 10 we report in the top line the results of the expected number of infected with the related confidence bands in case of homogeneous mixing and social mixing for the constant recovery rate *γ_e_*. At the bottom we compare the case of constant and age-dependent recovery rates for the social mixing scenario. In general, the homogeneous mixing hypothesis leads to an underestimation of the maximum number of infected and shows a slower decay over time, the latter effect also found in the presence of an age-dependent recovery rate. The expected total number of recovered people is shown in Figure 11. Finally in Figure 12 we report the expected age distribution of infectious individuals in time. We remind that all the simulations presented in this section have been obtained assuming to maintain the lockdown measures introduced on March 9 for all subsequent times.

**Figure 10:**
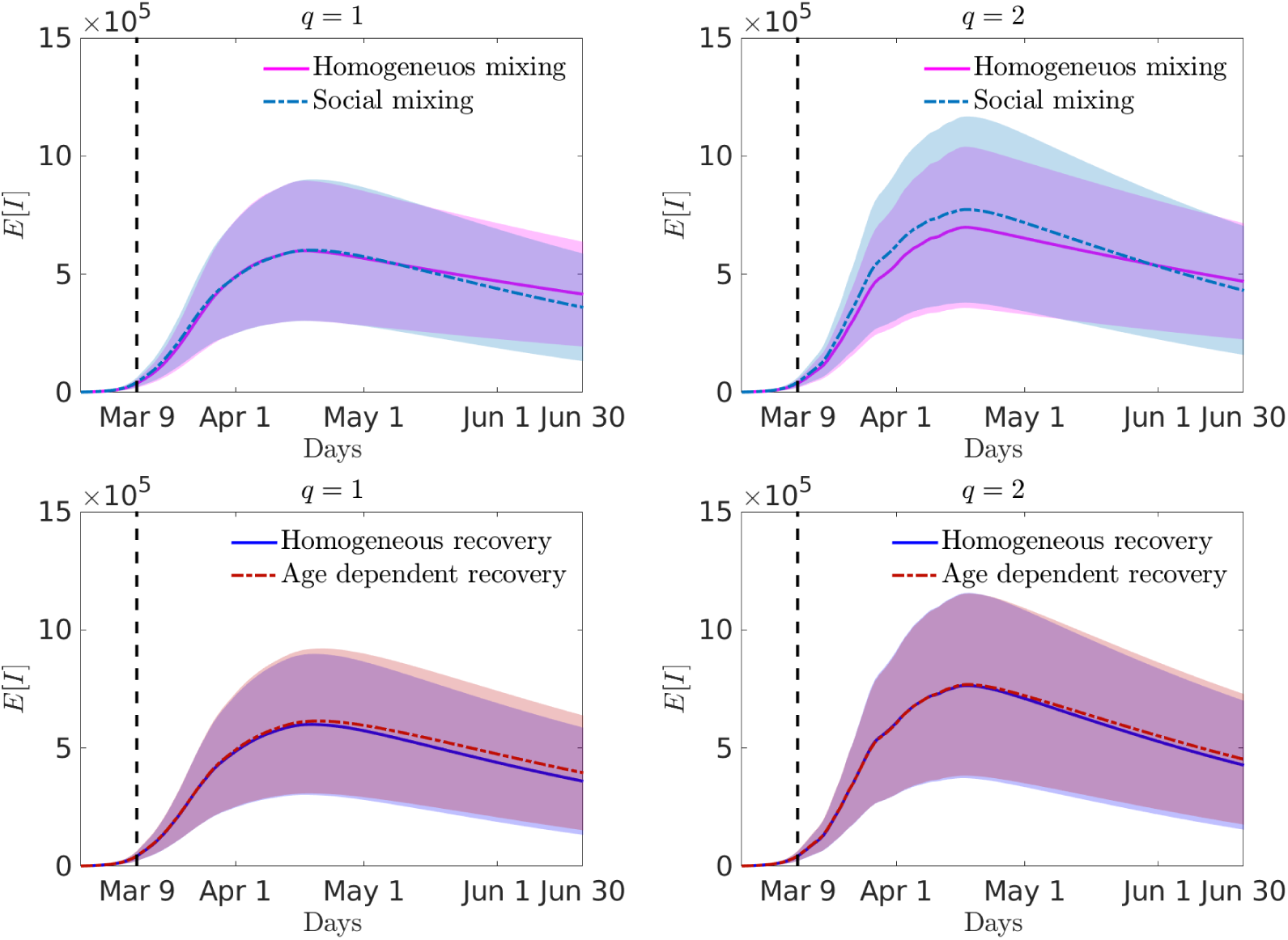
**Test 3**. Expected number of infected in time for *ψ*(*I*) = *I^q^*, *q* = 1 (left) and *q* = 2 (right) together with the confidence bands in the case of COVID-19 outbreak in Italy for homogeneous mixing or social mixing (top row) and, in the social mixing scenario, age-independent or age-dependent recovery rate (bottom row). The abscissae are measured in days starting from the beginning of the epidemic.

**Figure 11:**
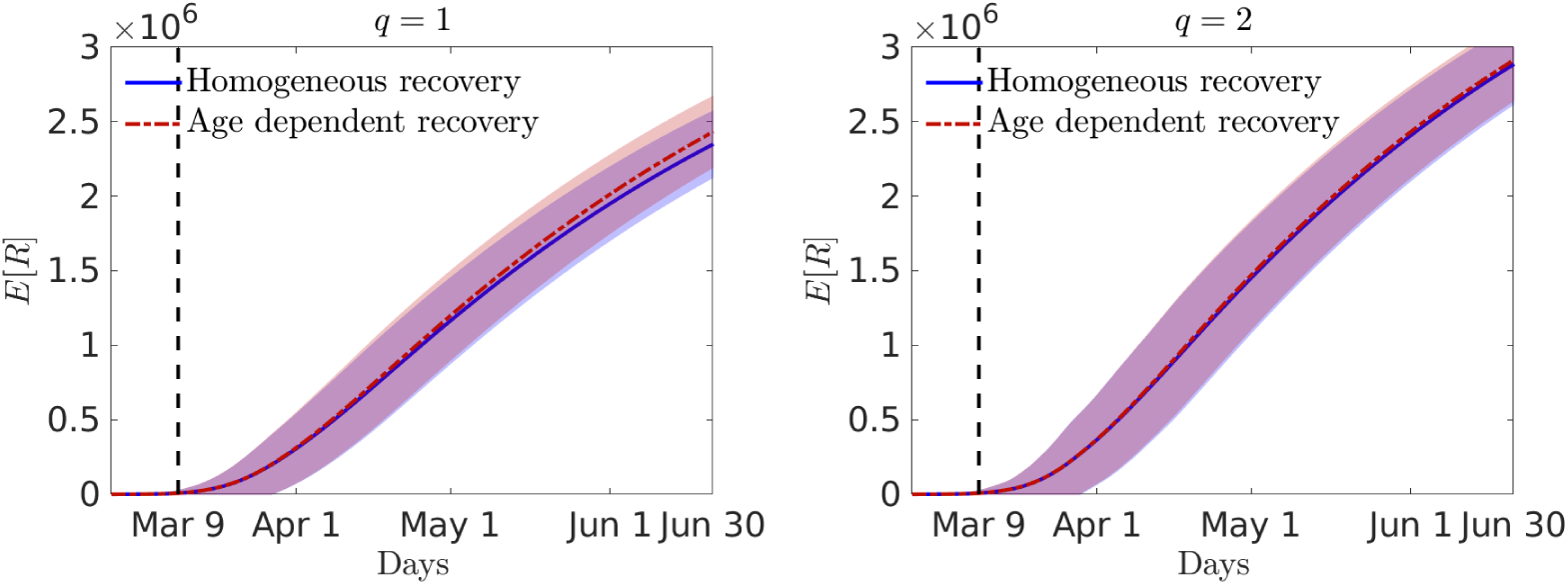
**Test 3**. Expected number of recovered in time for *ψ*(*I*) = *I^q^*, *q* = 1 (left) and *q* = 2 (right) together with the confidence bands in the case of COVID-19 outbreak in Italy for the social mixing scenario, age-independent or age-dependent recovery rate. The abscissae are measured in days starting from the beginning of the epidemic.

**Figure 12:**
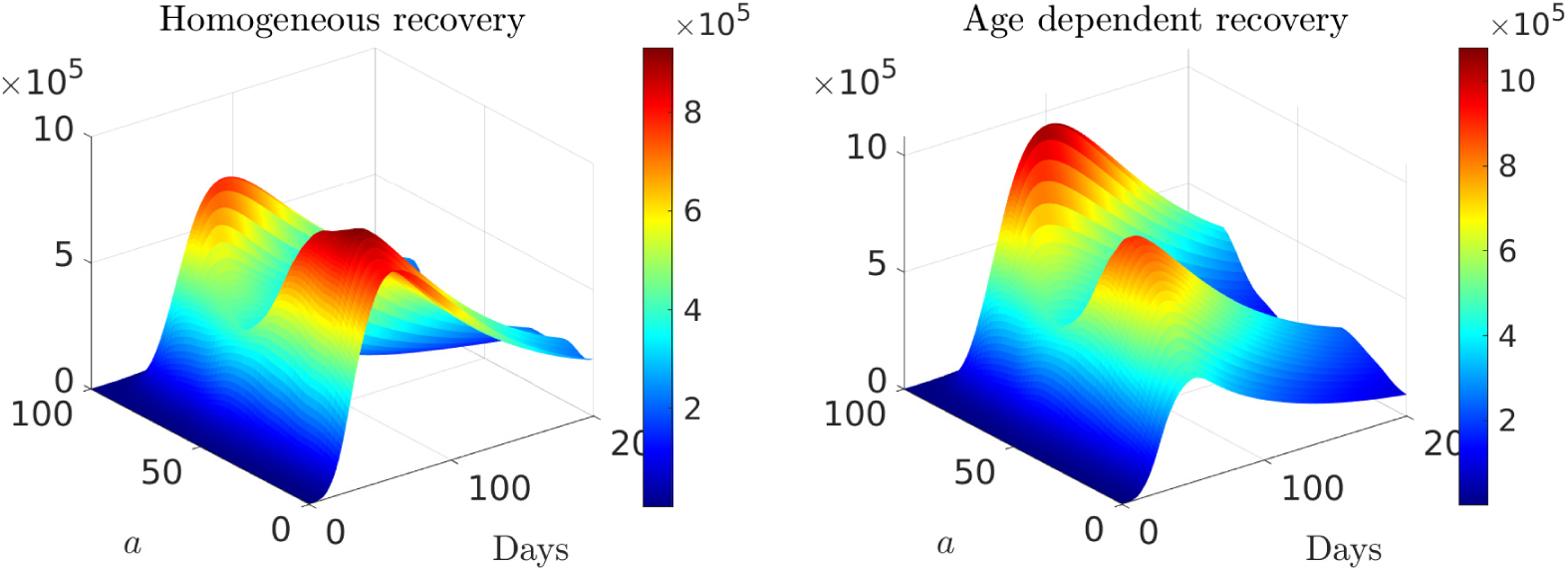
**Test 3**. Expected age distribution of infectious individuals over a long time horizon of 200 days in the social mixing scenario with *γ* = *γ_e_* (left) and age dependent recovery *γ*(*a*) defined in (33).

### 4.4 Test 4: Reducing the epidemic through relaxed social containment

One of the major problems in the application of very strong containment strategies, such as the lockdown applied in Italy, is the difficulty in maintaining them over a long period, both for the economic impact and for the impact on the population from a social point of view. In order to analyze sustainable control strategies, therefore, it is necessary to resort to models with a social structure and control methods based on specific forms of social distancing that allow the economy to restart and the population to dedicate itself, albeit in a limited way, to its pre-pandemic activities.

In accordance with the interaction function characterizing the productive activities introduced in the Appendix B, see also Figure 14, we considered the following age dependent penalization

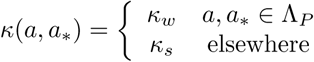

where Λ*_P_* defines the typical age group related to productive activities, for example Λ*_P_* = [20, 60] and 1*/κ_s_* > 1*/κ_w_* characterize two control actions related to a strong and a weaker containment of social distance. We will assume 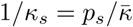 where 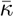 is the asymptotic value calculated for the penalization parameter in the lockdown period, while *κ_w_* has been relaxed compared to *κ_s_*, i.e.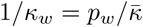, with 1 ≥ *p_s_* ≥ *p_w_*.

In Figure 13 we report the evolution of the age-controlled model in the case *q* = 1. We have considered two possible scenarios that reproduce the evolution of the infected in the hypothesis of relaxation of the lockdown measures at May 4 as foreseen by the Italian Government.

**Figure 13:**
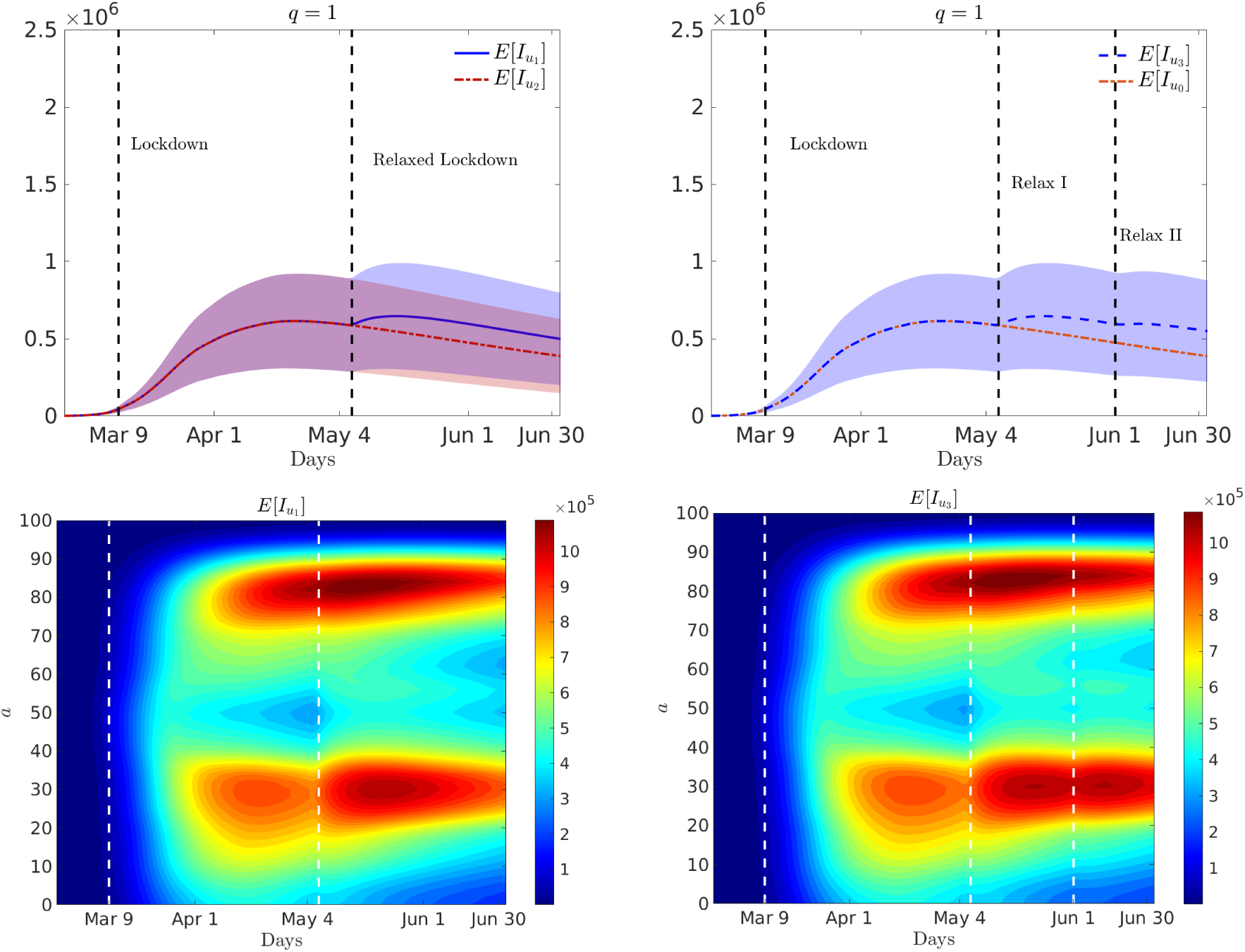
**Test 4**. Top row: evolution of the total number of infected in a two-zone scenario identified by *κ*_1_ and *κ*_2_ (left) and in a three zones scenario identified by *κ*_0_ and *κ*_3_. Bottom row: age distribution of infected in the two-zones (left) and three-zones (right) scenario.

In the first scenario, on the left, we consider two intervals that simulate the two different controlled periods measured in days from the beginning of the epidemic, 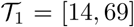 with *p_s_* = *p_w_* = 1, and 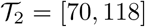 where the social measures of distance are loosened with *p_w_* = 0.6 and *p_s_* = 0.9. In Figure 13 (left) we compare this choice with a situation where the relaxation of the so-called phase 2 control measures in the interval 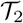 are further loosened by chosing *p_w_* = 0.3 and *p_s_* = 0.6. The respective numbers of infected in these two situations are denoted by 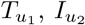. It is easily observed how we systematically increase the number of infected by successive relaxations of the initial strict social-distancing measures. In Figure 13 (bottom, left) we report also the evolution of age-dependent expected number of infected individuals.

The second scenario assumes that after an initial opening in which few productive activities are allowed, the government will gradually loosen the containment procedures with a gradual approach after a certain period of time. In this case we assumed three control zones: 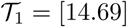 with *p_s_* = *p_w_* = 1, the interval 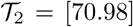 with *p_s_* = 0.9, *p_w_* = 0.6 and finally a third period 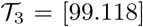 with *p_s_* = 0.8, *p_w_* = 0.5. This last period will correspond to a further relaxation of confinement measures on June 1st. The solution related to the number of infected is indicated with 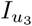 and is shown in Figure 13. (right). For comparison we also report the full lockdown solution denoted with 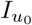. In the lower part of the figure on the right the corresponding distributions of infected individuals by age over time are also reported. As can be seen, a gradual strategy allows to contain the peak of contagions and to have a result comparable with the fully controlled model at a much lower social cost. The timing and intensity of the interventions, however, are crucial to prevent a restart of the epidemic wave.

## 5 Conclusions

Quantifying the impact of uncertain data in the context of an epidemic emergency is essential in order to design appropriate containment measures. Such containment measures, implemented by several countries in the course of the COVID-19 epidemic, have proved effective in reducing the *R*_0_ reproduction number to below or very close to one. These large-scale non-pharmaceutical interventions vary from country to country but include social distance (banning large mass events, closing public places and advising people not to socialize outside their families), closing borders, closing schools, measures to isolate symptomatic individuals and their contacts, and the large-scale lock-down of populations with all but essential prohibited travel.

One of the main problems is the sustainability of these interventions, which until the introduction of a vaccine will have to be maintained in the field for long periods. However, estimating the reproductive numbers of SARS-CoV-2 is a major challenge due to the high proportion of infections not detected by health care systems and differences in test application, resulting in diverse proportions of infections detected over time and between countries. Most countries currently have only the capacity to test a small proportion of suspected cases and tests are reserved for severely ill patients or high risk groups. The available data therefore offer a systematic partial overview of trends.

In this article, starting from a SIR-type compartmental model with social structure, we have developed new mathematical models describing the actions of a government agency to control the population in order to reduce the estimated number of infected people in the presence of uncertain data. This hypothesis allows to derive a model that contains the control action in feedback form based on the perception of the policy maker of the spread of the disease. Subsequently the model has been effectively solved in the presence of uncertain data, expanding the state variables into orthogonal polynomials in the uncertainty space, reducing the problem to a set of deterministic equations for the distribution of the solution through the course of the epidemic. The resulting controlled dynamic system then results in a deterministic stochastic solution that enables efficient estimation of uncertain parameters.

The numerical simulations, carried out using data from the recent COVID-19 outbreak in Italy, show, on the one hand, the model’s ability to well describe lockdown scenarios aimed at flattening the infection curve and, on the other hand, how the high uncertainty of the data on the number of infected people makes it very difficult to provide long-term quantitative forecasts. By identifying some plausible scenarios in accordance with the literature, sustainable containment measures by the population based on the resumption of certain occupational activities characterized by specific age groups and social interaction matrices have been studied. The results demonstrate the effectiveness of such approaches based on the social structure of the system capable of achieving results similar to much more restrictive policies in reducing the risk that the virus may return to spread when the restrictions are lifted but at a significantly lower socio-economical cost.

Further studies in this direction will be aimed at considering more realistic epidemic models than the one analyzed in this work together with multiple control terms specific for each social activity in order to design optimal strategies to mitigate the overall epidemic impact.

## Data Availability

All the data referred to in the manuscript are publicly available.

## Acknowledgements

G. A. and L. P. acknowledge the support of PRIN Project 2017, No. 2017KKJP4X “Innovative numerical methods for evolutionary partial differential equations and applications”. This research was partially supported by the Italian Ministry of Education, University and Research (MIUR) through the “Dipartimenti di Eccellenza” Programme (2018–2022) – Department of Mathematics “F. Casorati”, University of Pavia. M.Z. is member of GNFM (Gruppo Nazionale per la Fisica Matematica) of INdAM (Istituto Nazionale di Alta Matematica), Italy.

### A Stochastic Galerkin approximation

In this Appendix we give the details of the stochastic Galerkin (sG) method used to solve the feedback controlled system (25)-(26) with uncertainties. To this aim, we consider a random vector **z** = (*z*_1_, *…,z_d_*) with independent components and whose distribution is 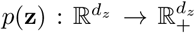 The stochastic Galerkin approximation of the differential model (25)-(26) is based on stochastic orthogonal polynomials and provides a spectrally accurate solution under suitable regularity assumptions, see [40]. We consider the linear space ℙ*_M_* of polynomials of degree up to *M* generated by a family of polynomials 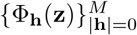 that are orthonormal in the space *L*^2^(Ω) such that

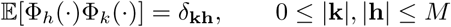

being **k** = (*k*_1_, *…,k_d_*) a multi-index, 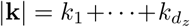 with *δ_kh_* the Kronecker delta function, and 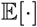 the expectation with respect to *p*(**z**). The construction of the polynomial basis 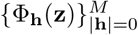 depends on the distribution of the uncertainties and must be chosen in agreement with the Askey scheme [40]. We summarise in Table 1 several polynomials bases in connection with the law of a random component of **z**.

**Table 1:** The different polynomial expansions connected to the probability distribution of the random component *z_k_*, *k* = 1,…,*d_z_*.

| Probability law | Expansion polynomials | Support |
| --- | --- | --- |
| Gaussian | Hermite | $(-\infty, +\infty)$ |
| Uniform | Legendre | $[a, b]$ |
| Beta | Jacobi | $[a, b]$ |
| Gamma | Laguerre | $[0, +\infty)$ |
| Poisson | Charlier | $\mathbb{N}$ |

Assuming now *s*(**z**, *a, t*), *i*(**z**, *a, t*) and *r*(**z**, *a, t*) in *L*^2^(Ω) we may approximate these terms through a generalized polynomial chaos expansion in the random space as follows

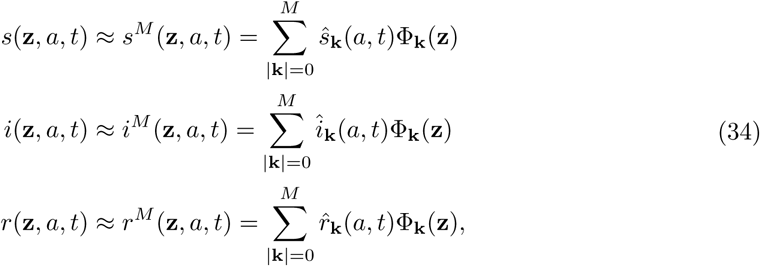

where the quantities 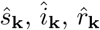 are projections in the polynomial space

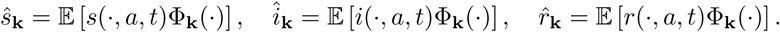

The sG formulation of system (25) is obtained first by replacing the solutions with their stochastic polynomial expansions

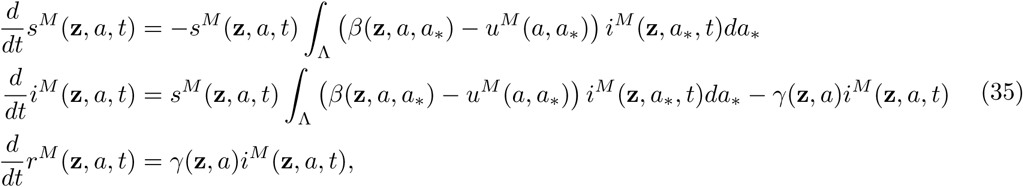

with

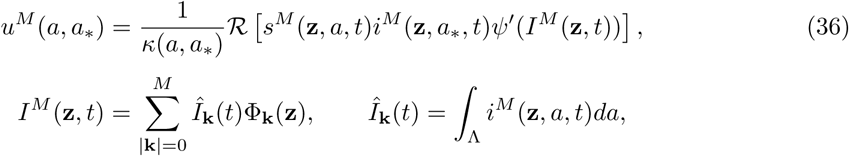

and where *s^M^*, *i^M^*, *r^M^*, rare defined by (34). Then, thanks to the orthonormality of the polynomial basis of ℙ*_M_*, multiplying by Φ_m_, for all |**m**|≤ *M*, and taking the expectation with respect to *p*(**z**) we obtain the following coupled system of *M* + 1 deterministic equations for the evolution of each projection

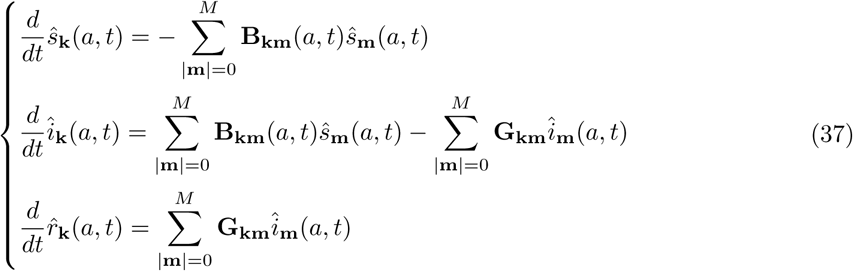

where

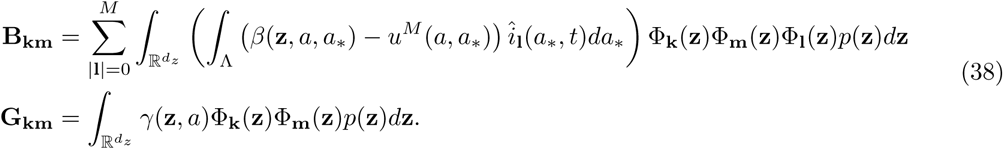

The above system is then integrated in time directly in the space of projections. We remark that statistical quantities of interest, such as expectation and variance of infected, can be recovered as

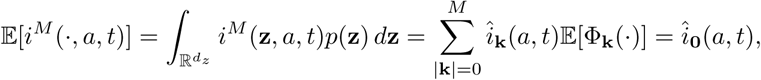

whereas for the variance we get

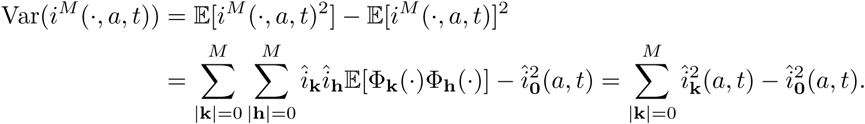

### B Social mixing functions

In this appendix we report the details of the social interaction functions that characterize the dynamics of social mixing. These characteristics are in fact crucial for a correct prediction of outcomes, especially in diseases transmitted by close contacts. Several large-scale studies have been designed in the last decade to determine relevant age-based models in social mixing. Without attempting to review the vast literature on this topic, we mention [6, 33, 35] and the references therein.

The number of contacts per person generally shows considerable variability according to age, occupation, country and even day of the week, in relation to the social habits of the population. Nevertheless, some universal behaviours can be extracted which emerge as a function of specific social activities. Social mixing is highly age-related, which means that people usually tend to interact with other people of a similar age. Young people have a high rate of contact with adults aged around 30–39 and older people over 60, i.e. their parents and grandparents respectively. Contact rates are indeed very high at home and at school. On the other hand, professional mixing is weakly assortative by age and tends to be determined by uniform interactions, approximately between people from 25 and 60 years old.

For these reasons we consider an interaction function determined by three main sub-functions that characterize the family, the school and the professional mixing. Therefore, a stylized function approximating a realistic contact matrix can be written as follows

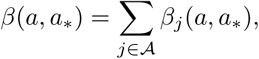

where the functions *β_j_*(*a, a*_*_) take into account the different contact rates of people with ages *a* and *a*_*_ in relation to specific social activities of the type 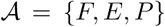, where we identify family contacts with *F*, education and school contacts with *E* and professional contacts with *P*. The particular structure of these social interaction matrices was determined empirically in [6,35]. Here, according to these observations, we propose suitable mathematical functions that can be calibrated to reproduce empirical observations.

In details, familiar contacts tend to concentrate on a three-band matrix with a peak around younger ages. This can be reproduced considering the function

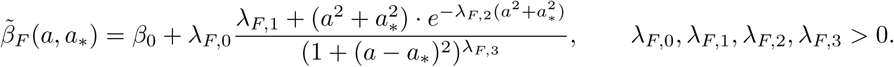

Hence, we define the family interactions as

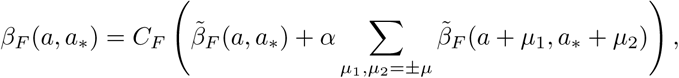

being *µ* > 0 the age shift at which family contacts occur.

On the other hand, school and professional interactions are more age-specific and the corresponding matrices can be reproduced as follows

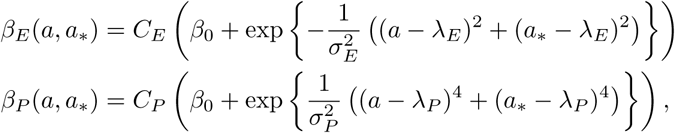

being *C_F_, C_E_, C_P_* > 0 normalizing constants such that *∫*_Λ_*β*(*a, a*_*_)*da da*_*_ = 1, λ*_E_* > 0 is the average contact age at school, and λ*_P_* > 0 is the average professional contact age. In Figures 14 and 15 we represent the three social interaction functions and the resulting global social interaction function *β*(*a, a*_*_). The details of the parameters used in the simulations are reported in Table 2.

**Figure 14:**
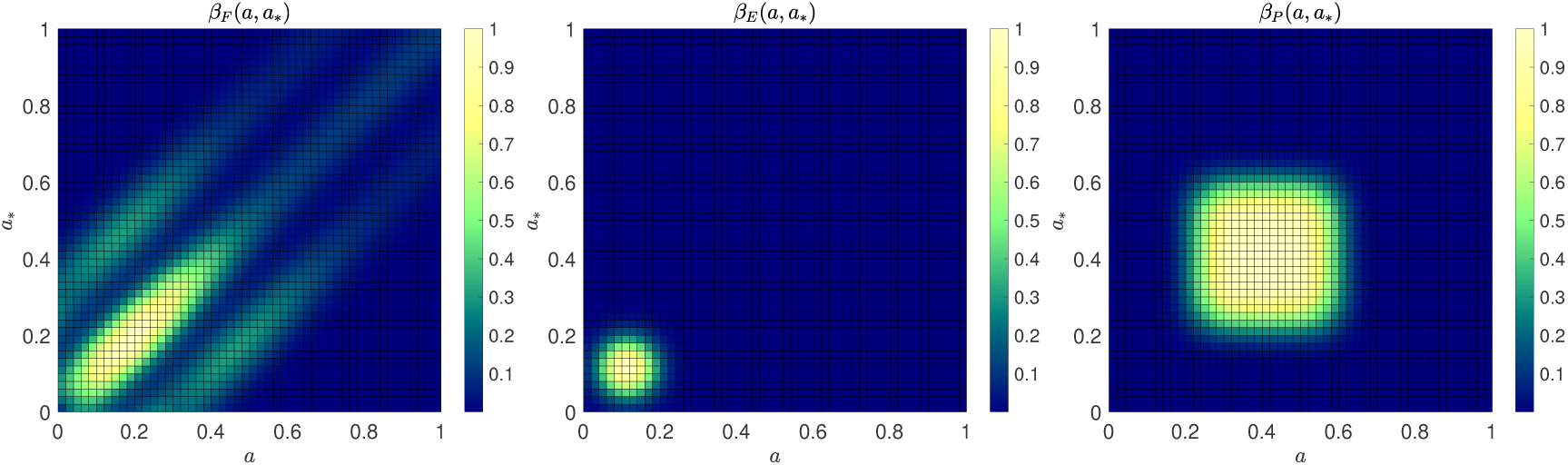
From left to right, contour plot of the social interaction functions *β_F_*, *β_E_* and *β_P_* taking into account the different contact rates of people with ages *a* and *a*_*_ in relation to specific social activities. The function *β_F_* characterizes the family contacts, *β_E_* the education and school contacts, and *β_P_* the professional contacts.

**Figure 15:**
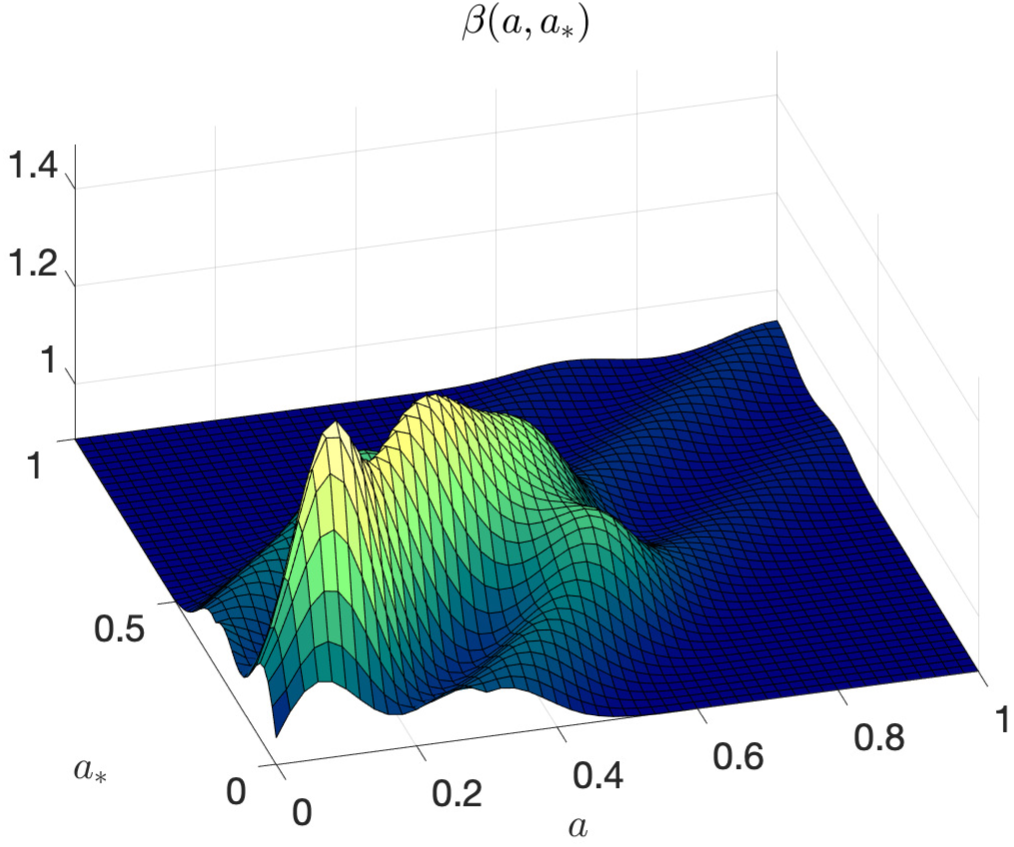
The social interaction function *β* = *β_F_* + *β_E_* + *β_P_*.

**Table 2:** The parameters defining the details of the interaction functions used in the simulations.

| Contact function | Parameters |  |  |  |  |  |  |
| --- | --- | --- | --- | --- | --- | --- | --- |
| $\beta_F(a, a_*)$ | $\beta_0$ | $\lambda_{F,0}$ | $\lambda_{F,1}$ | $\lambda_{F,2}$ | $\lambda_{F,3}$ | $\mu$ | $\alpha$ |
|  | 4/3 | 2 | 1/100 | 8 | 100 | 3/10 | 1/4 |
| $\beta_E(a, a_*)$ | $\beta_0$ | $\sigma_E^2$ | $\lambda_E$ | | | | |
|  | 4/3 | 1/20 | 21/20 |  |  |  |  |
| $\beta_P(a, a_*)$ | $\beta_0$ | $\sigma_P^2$ | $\lambda_P$ | | | | |
|  | 4/3 | 1/40 | 2/5 |  |  |  |  |

## Footnotes

1 Q&A: Similarities and differences – COVID-19 and influenza. https://www.who.int/news-room/q-a-detail/q-a-similarities-and-differences-covid-19-and-influenza

2 Source ISTAT (https://www.istat.it) and Istituto Superiore di Sanità (https://www.epicentro.iss.it)

